# Center-of-Mass Work Patterns Reveal a Dissociation Between Gait Organization and Limb-level Mechanical Function in Post-stroke Walking

**DOI:** 10.64898/2026.04.14.26350877

**Authors:** Seyed-Saleh Hosseini-Yazdi, Karson Fitzsimons, John EA Bertram

## Abstract

Walking speed is widely used to assess gait recovery following stroke, yet it provides limited insight into how walking performance is mechanically organized. This study examined how center-of-mass (COM) work organization and propulsion–support coupling vary across walking speeds in individuals with post-stroke hemiparesis to distinguish recovery of gait organization from recovery of limb-level mechanical function. Eleven individuals with post-stroke hemiparesis performed treadmill walking across speeds ranging from 0.2 to 0.7 m/s while ground-reaction forces were recorded. Limb-specific COM power and work were computed using an individual-limbs framework, and interlimb asymmetry in net and positive work, along with the propulsion–support ratio (PSR), were quantified. A qualitative transition in gait organization was observed: at lower walking speeds, COM power exhibited a simplified two-phase pattern, whereas at higher walking speeds (approximately ≥0.5 m/s), a structured four-phase COM power pattern emerged, including identifiable push-off and preload phases. Despite this recovery of gait organization, interlimb work asymmetry remained elevated and paretic PSR remained reduced across all speeds, indicating persistent limb-level mechanical deficits. These findings demonstrate that increases in walking speed and the emergence of typical COM power structure reflect recovery of gait organization rather than restoration of underlying limb-level mechanical capacity. Consequently, walking speed alone is insufficient to characterize gait recovery after stroke, and biomechanically informed measures of COM work organization and propulsion–support coupling provide complementary insight by distinguishing organizational recovery from limb-level mechanical recovery.

## Introduction

Walking speed is the most widely used measure of gait recovery following stroke[1–4], owing to its simplicity and strong association with functional outcomes[5]. Speed-based classifications [6–9] are routinely used to define impairment severity [2] and track rehabilitation progress [9,10]. However, walking speed is an outcome descriptor rather than a mechanistic measure [6,11]: it quantifies how fast an individual walks, but not how gait is mechanically organized to achieve that performance [6,12].

A fundamental challenge in interpreting post-stroke gait is that similar walking speeds can arise from markedly different mechanical strategies [13,14]. In hemiparetic gait, increases in walking speed often co-occur with persistent deficits [15,16], including reduced paretic propulsion [12,17,18], asymmetric ground-reaction forces [19–24], and prolonged double support [13,25– 27]. These features are well established, yet their interpretation remains ambiguous. In particular, gains in speed are frequently taken as evidence of recovery [4,13,26,28–31], despite the possibility that they arise alongside unresolved deficits in limb-level mechanical function. Critically, it remains unclear whether recovery of gait organization and recovery of limb-level mechanical function occur concurrently or can be mechanistically dissociated in post-stroke walking. As a result, walking speed alone may overestimate recovery by obscuring persistent asymmetries in how mechanical work is generated and distributed between limbs [32,33].

A complementary perspective is to consider walking as a mechanically coordinated process in which the limbs generate and redistribute work to support and redirect the body’s center of mass (COM) across successive steps [34,35]. From this viewpoint, gait is defined by how propulsion and support are produced and coordinated at the whole-body level [36]. Measures derived from COM power and work provide a direct description of this organization, capturing limb-specific contributions to step-to-step transitions [37–39] as well as the coupling between vertical support and forward progression [40,41]. Although such measures have been used to characterize gait impairment [42,43], their potential to reveal changes in gait organization across walking speeds remains underexplored in post-stroke populations.

In neurologically intact walking, systematic changes in COM power structure and limb-level work organization are observed across walking speeds [44]. In post-stroke hemiparetic gait, however, it remains unclear whether walking at higher speeds reflects a continuous scaling of impaired mechanics or a qualitative reorganization of gait management [45]. In particular, the emergence of structured features characteristic of typical walking—such as identifiable push-off and multi-phase COM power patterns—may be interpreted as evidence of recovery. Whether these features reflect recovery of gait organization, recovery of paretic limb mechanical function, or a combination of both remains unexplored.

The present study addresses this gap by characterizing how COM work organization and propulsion–support coupling vary across walking speeds in individuals with post-stroke hemiparesis. Using ground-reaction-force data and an individual-limbs framework[34,36], we quantify limb-specific COM power and work and derive measures of interlimb work asymmetry and propulsion–support coupling. We hypothesize that adopting higher walking speeds is associated with a qualitative transition in gait organization, marked by the emergence of structured COM power patterns, while underlying limb-level mechanical deficits persist. By distinguishing recovery of gait organization from recovery of mechanical capacity, this study provides a mechanistic framework for interpreting speed-dependent gait behavior in post-stroke walking.

## Materials and Methods

This study employed a repeated-measures observational design to examine gait mechanics in individuals with post-stroke hemiparesis across a range of walking speeds. Data were collected during treadmill walking sessions conducted as part of participants’ routine inpatient rehabilitation program. No experimental interventions were introduced, and all walking activities were performed under standard clinical supervision.

### Participants

Participants were recruited from the Neuro Rehabilitation Unit at Foothills Hospital in Calgary, Canada. Inclusion criteria were: (1) age ≥18 years; (2) first-time ischemic or hemorrhagic stroke within the previous 6 months; and (3) ability to walk with or without assistive devices. Exclusion criteria included: (1) additional neurological disorders; (2) significant lower-limb orthopedic impairments; (3) uncontrolled cardiovascular conditions; or (4) cerebellar stroke.

Eleven participants (5 female, 6 male) were included in the study. Mean (SD) age was 54.6 (11.5) years, and mean (SD) time post-stroke at first assessment was 10.0 (7.0) weeks. All participants provided written informed consent prior to participation. The study protocol was approved by the Conjoint Health Research Ethics Board at the University of Calgary (REB21-1576).

### Clinical Assessment

Baseline clinical assessments were completed as part of standard inpatient evaluation. Overground self-selected walking speed was assessed using the average of two 10-m walk tests. Walking endurance was evaluated using the 6-minute walk test (6MWT), and lower-extremity motor impairment was quantified using the Fugl-Meyer Lower Extremity (FM-LE) assessment. Participants were permitted to use their usual assistive devices (e.g., cane or walker) during all overground clinical assessments.

### Experimental Protocol

Participants completed multiple treadmill walking sessions (up to three sessions per week) during their inpatient rehabilitation stay. Each session included at least 2 minutes of walking on a split-belt instrumented treadmill (Bertec Corp., Columbus, OH, USA) with embedded force plates recording ground reaction forces (GRFs) at 1000 Hz.

Treadmill speed was selected and progressively adjusted by the supervising physiotherapist based on each participant’s functional capacity and clinical judgment. Participants wore a safety harness capable of fall arrest and were permitted light hand contact with a front handrail if required for balance. A semi-immersive visual environment was provided, with visual flow matched to treadmill speed. Heart rate and blood pressure were monitored before and after each walking bout as part of routine clinical safety procedures.

### Data Processing

GRF data were low-pass filtered using a third-order Butterworth filter (cutoff frequency: 10 Hz) with zero-phase lag. Center-of-mass (COM) velocity was computed from the GRFs using standard numerical integration. To minimize integration drift, COM velocity signals were high-pass filtered at 0.25 Hz[34,46]. Limb-specific COM power was calculated using the individual-limbs method, which decomposes total COM power into contributions from each limb and resolves simultaneous positive and negative work performed during double support[36,47].

### COM Work Analysis

COM power profiles were used to quantify mechanical work performed during walking. When a distinct four-phase structure—collision, rebound, preload, and push-off—was identifiable, phase-specific work was computed by time integration of COM power within each phase. When this structure was not clearly present, COM work was summarized as total positive and total negative work per step[36,48] (Figure *1*).

**Figure 1:**
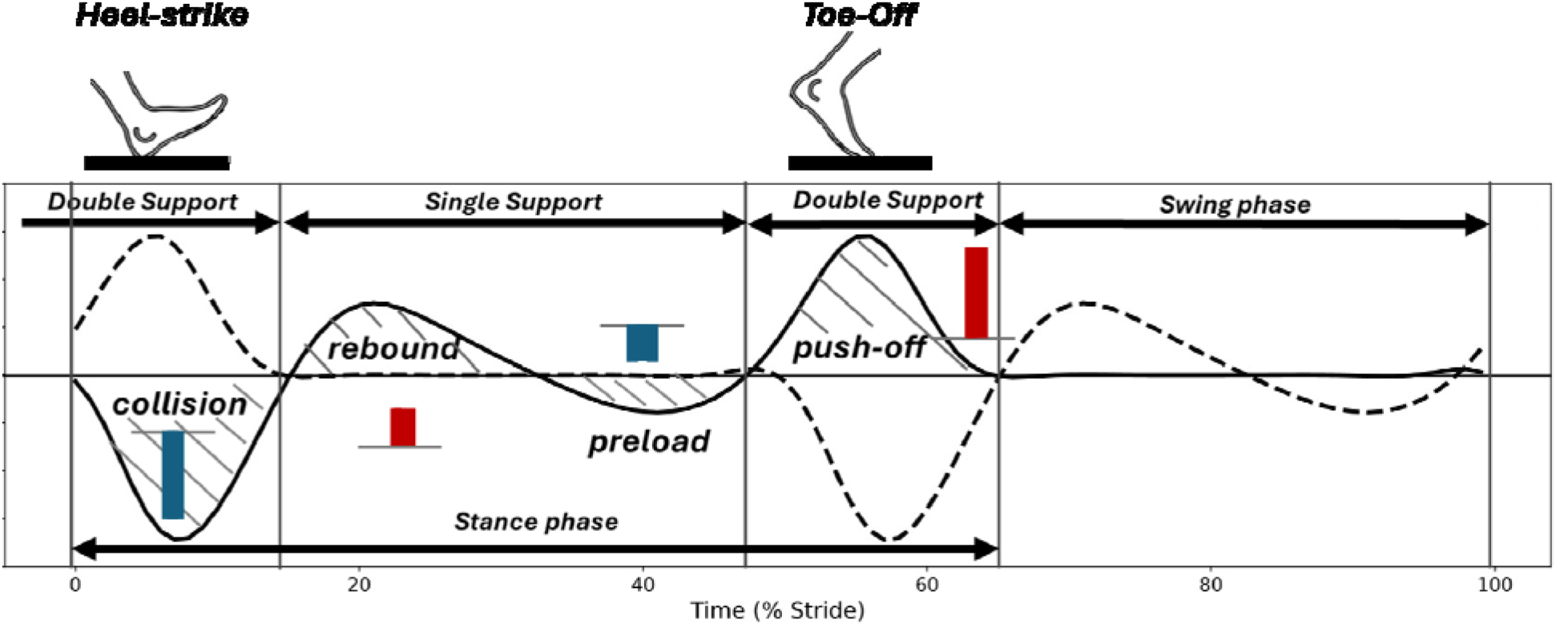
It is shown that in healthy nominal walking, the center of mass (COM) power depicts four distinct phases: heel strike collision, rebound, preload, and push-off (adopted from [*48*]). The associated work of each phase is calculated by tim integration of the associated power profile. Negative work is indicated by dark blue columns while positive work are indicated by dark red columns.

### Outcome Measures

#### Interlimb COM Work Asymmetry

Net COM work asymmetry was defined as:

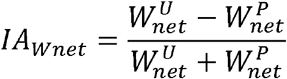

where 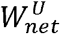 and 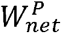 represent net COM work performed by the unaffected and paretic limbs, respectively. This metric quantifies the imbalance in overall mechanical work generation between limbs, with higher values indicating greater reliance on the non-paretic limb.

Positive work asymmetry was similarly defined as:

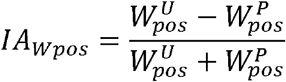

where 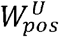 and 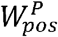 denote positive COM work generated by the unaffected and paretic limbs, respectively. This metric reflects asymmetry in propulsion-related work.

#### Propulsion–Support Ratio (PSR)

The propulsion–support ratio (PSR) was defined as:

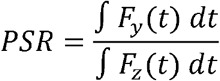

where *F*_*y*_ (*t*) is the anterior–posterior GRF and *F*_*z*_ (*t*) is the vertical GRF. PSR quantifies how effectively vertical load support is converted into forward propulsion.

### Speed Stratification

Walking trials were grouped into five discrete speed bins (0.2, 0.3, 0.4, 0.5, and 0.7 m/s). For each participant, data from multiple sessions were pooled within each speed bin to characterize speed-dependent changes in gait mechanics.

### Statistical Analysis

Descriptive statistics are reported as mean ± standard deviation. Relationships between walking speed and biomechanical outcome measures were evaluated using regression analyses. Piecewise linear regression was used to assess potential changes in trends between lower and higher walking speeds, and quadratic regression models were used to evaluate nonlinear relationships. Statistical significance was set at *p* < 0.05.

## Results

### Ground Reaction Forces and COM Power Profiles

At lower walking speeds (0.2–0.4 m/s), vertical ground reaction force (GRF) profiles did not exhibit the characteristic double-peaked pattern associated with typical walking. Peak vertical forces were consistently lower on the paretic limb compared to the non-paretic limb. In the anterior–posterior direction, the paretic limb generated relatively larger braking impulses, whereas the non-paretic limb generated larger propulsive impulses (Figure *2*).

**Figure 2:**
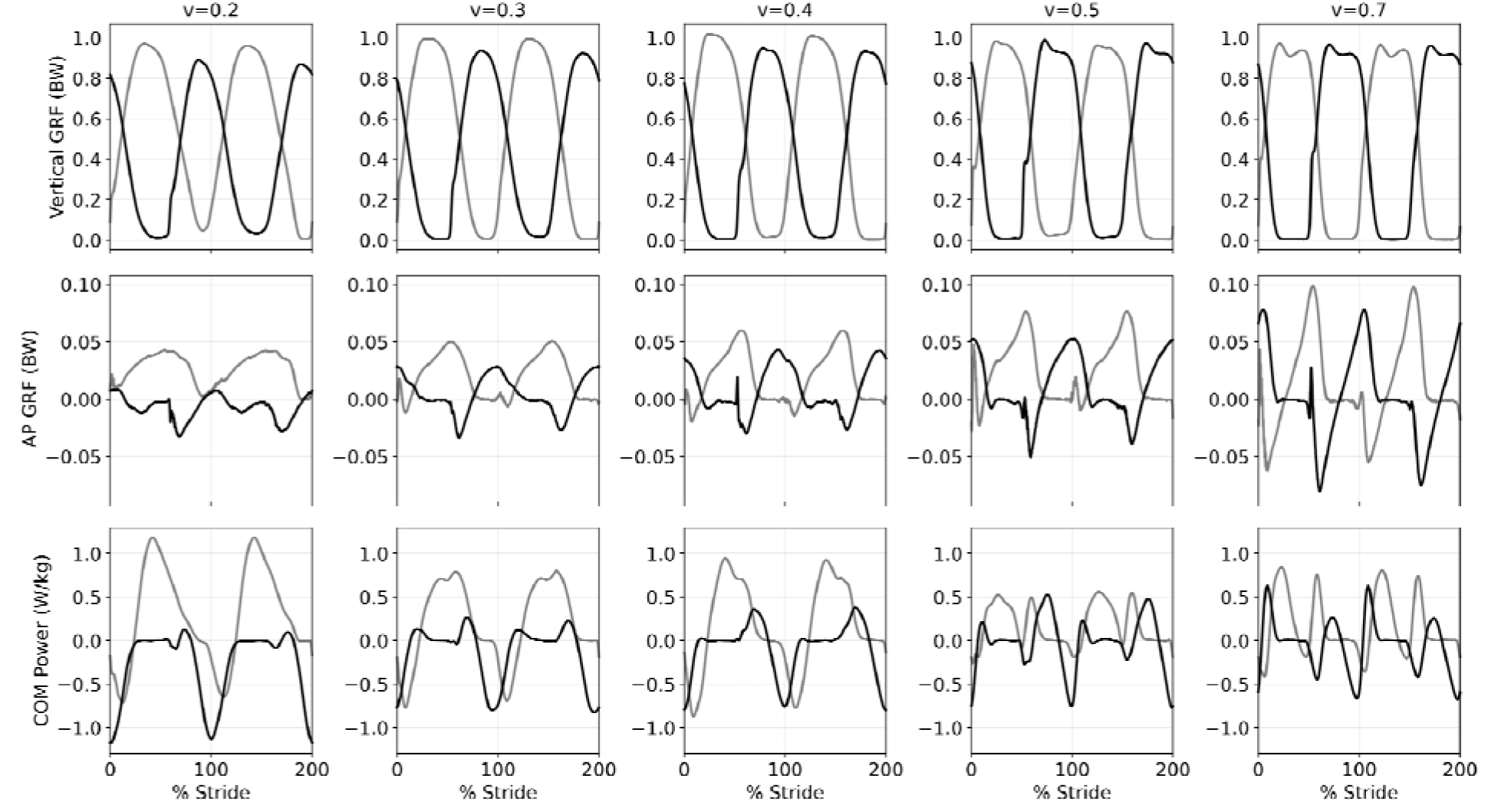
The hemi-paretic walking data collection based on walking speed: 0.2, 0.3, 0.4, 0.5 and 0.7. The top row shows the average vertical ground reaction force, the middle row is for the average PA ground reaction force, and the bottom row is for the average COM power. For speeds less than 0.5, the COM power profile shows a two-phase trajectory. The four-phase trajectory emerges for walking speeds ≥ 0.5. The paretic traces are shown in black while unaffected sides are depicted in gray.

Correspondingly, center-of-mass (COM) power profiles at these lower speeds exhibited a simplified structure characterized by alternating positive and negative work phases. A distinct four-phase pattern—corresponding to collision, rebound, preload, and push-off—was not observed within this speed range.

At higher walking speeds (≥0.5 m/s), vertical GRF profiles began to show a more defined double-peaked structure. Concurrently, COM power profiles demonstrated the emergence of a four-phase pattern, including identifiable push-off during late stance (Figure *2*). This transition was accompanied by the appearance of positive work performed by the paretic limb during late stance.

### COM Work Organization Across Speeds

At walking speeds below 0.5 m/s, COM work was characterized by two dominant components: total positive work and total negative work per step. In this range, work between limbs alternated without clear separation of preload and push-off phases (Figure *3*).

**Figure 3:**
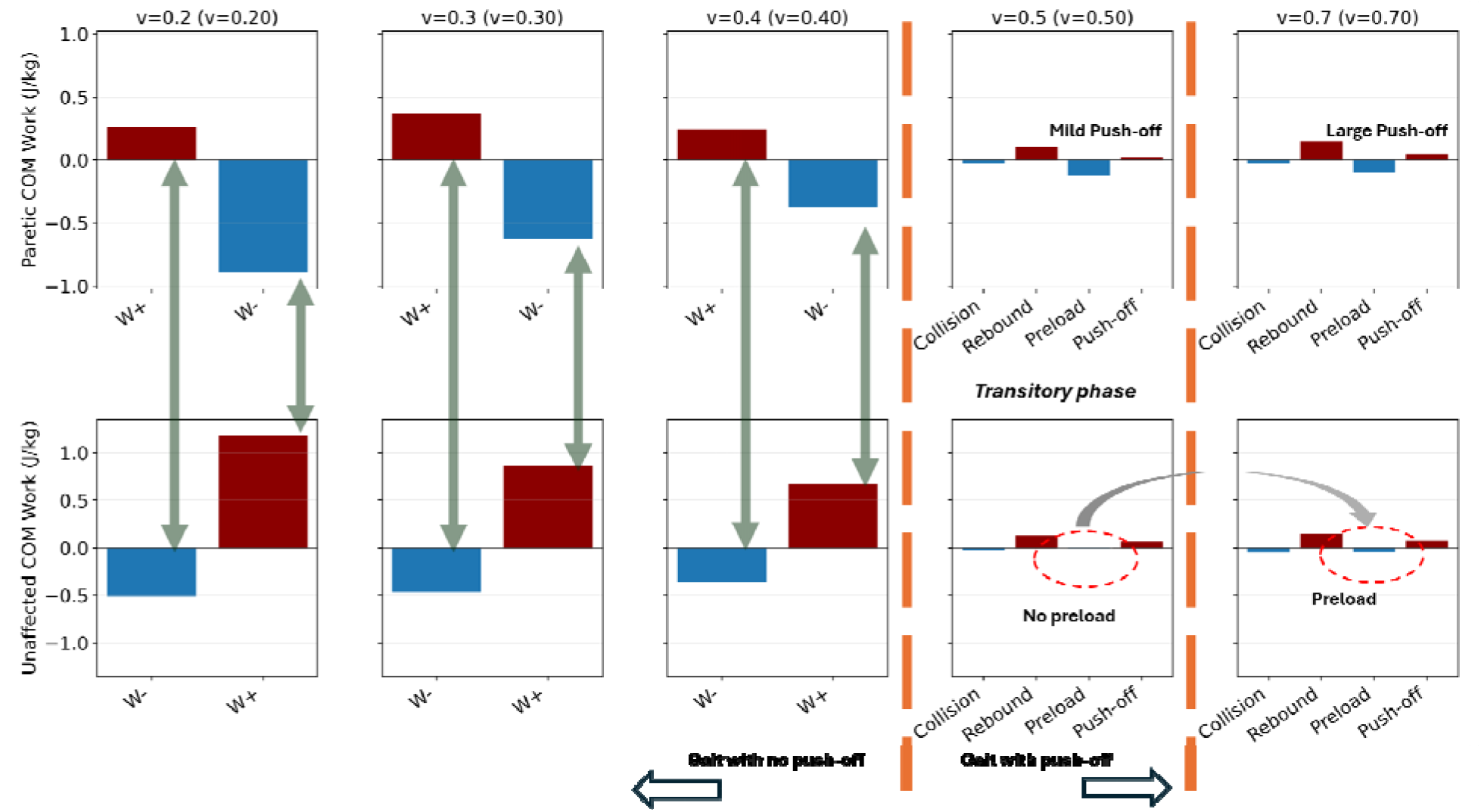
The COM work components of hemi-paretic walking: the top row indicates the paretic side while the bottom row represents the unaffected side. We present the positive work as dark red whereas blue indicates step negative work. For 0.2, 0.3, and 0.4 where the push-off is not detected, there are only two dominant work phases: positive and negative. At 0.5 paretic push-off appears and both sides show four-phase COM power profiles. At 0.5, single support preload is negligible, yet its magnitude rises at 0.7.

At walking speeds of 0.5 m/s and above, COM work organization became more structured. Phase-specific components, including rebound, preload, and push-off, were identifiable. Paretic limb push-off work was minimal or absent at lower speeds but became detectable at 0.5 m/s and increased further at 0.7 m/s. Single-support preload work remained small at 0.5 m/s and increased at higher speeds.

### Interlimb COM Work Asymmetry

Net COM work asymmetry was elevated across all walking speeds. Mean (SD) values were 0.783 (0.582) at 0.2 m/s, 0.846 (0.489) at 0.3 m/s, and 1.000 (0.174) at 0.4 m/s. At higher speeds, asymmetry decreased to 0.684 (0.428) at 0.5 m/s and increased again to 0.822 (0.262) at 0.7 m/s.

Piecewise regression analysis indicated a positive trend in net work asymmetry at lower speeds (slope = 1.28, p = 0.053, R^2^ = 0.10) and no significant association at higher speeds (slope = 0.69, p = 0.48, R^2^ = 0.04) (Figure *4*A).

Positive COM work asymmetry demonstrated a nonlinear relationship with walking speed. Mean (SD) values were 103.38% (66.42) at 0.2 m/s, 75.54% (52.49) at 0.3 m/s, and 79.92% (44.23) at 0.4 m/s. Asymmetry decreased to 25.26% (28.47) at 0.5 m/s and increased to 55.24% (45.51) at 0.7 m/s.

Quadratic regression analysis revealed a significant relationship between walking speed and positive COM work asymmetry (β_2_ = 642.89, p = 0.023, R^2^ = 0.23) (Figure *4*B).

**Figure 4:**
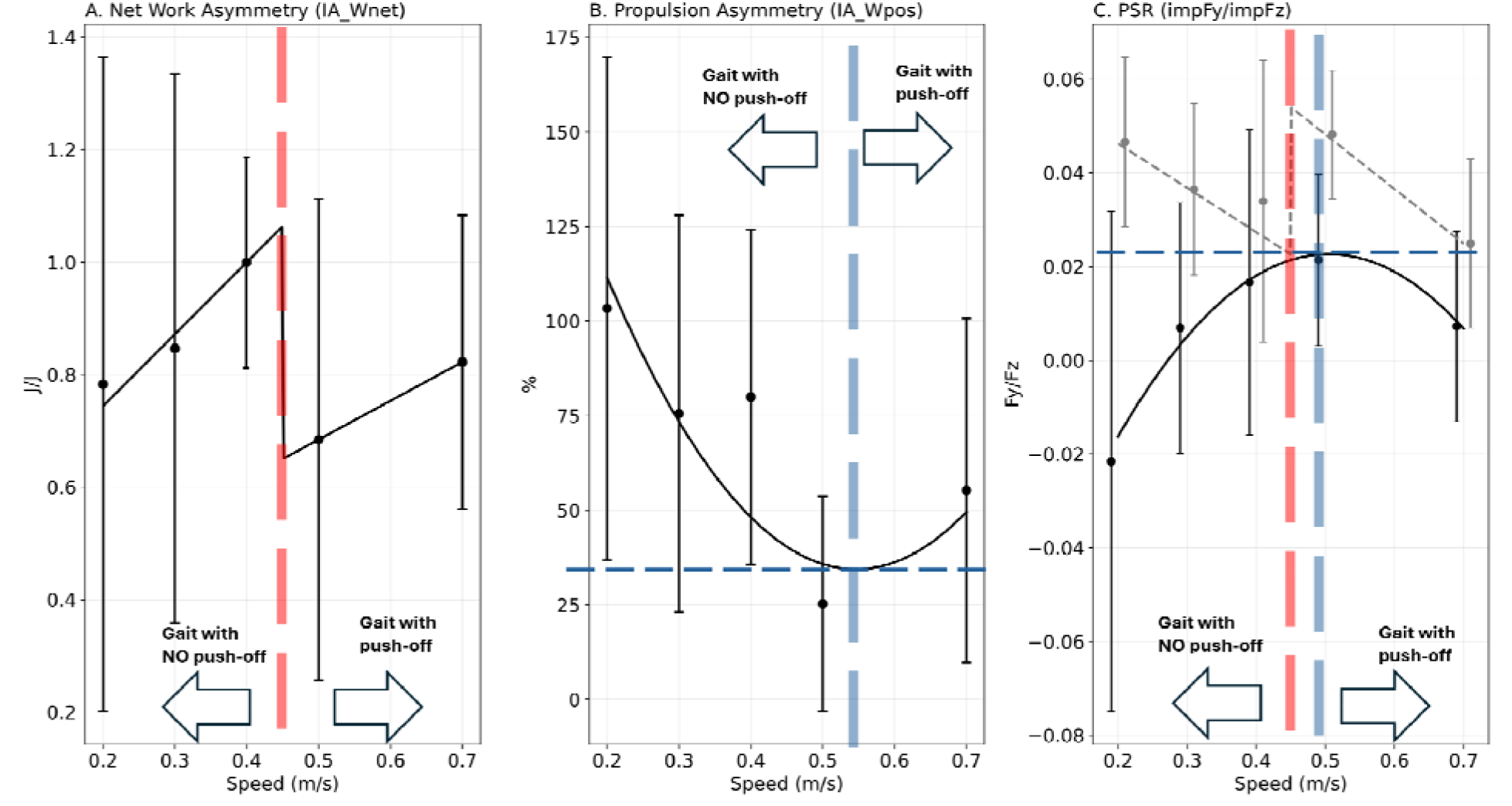
Hemiparetic walking asymmetry evaluation indices: (A) Net work asymmetry index measuring the asymmetry in net work performance of paretic and unaffected side, (B) the Propulsion index of asymmetry representing the asymmetry in positiv work of either side, and (C) the PSR index for paretic and unaffected sides. All indices suggest a transition in the state of hemiparetic walking around 0.5. The solid line represents the PSR for the paretic side while the dashed gray lin show the unaffected side. The detected transitions are also indicated by vertical dashed lines.

### Propulsion–Support Ratio (PSR)

The paretic limb propulsion–support ratio (PSR) was low across all walking speeds. Mean (SD) values were −0.0216 (0.0533) at 0.2 m/s, 0.0069 (0.0267) at 0.3 m/s, and 0.0166 (0.0325) at 0.4 m/s. At higher speeds, paretic PSR increased slightly to 0.0214 (0.0183) at 0.5 m/s and decreased to 0.0072 (0.0204) at 0.7 m/s.

Quadratic regression analysis demonstrated a significant nonlinear relationship between walking speed and paretic limb PSR (β_2_ = 0.419, p = 0.012, R^2^ = 0.14).

In contrast, the non-paretic limb PSR remained consistently higher across all walking speeds. Mean (SD) values were 0.0466 (0.0181) at 0.2 m/s, 0.0365 (0.0183) at 0.3 m/s, 0.0340 (0.0301) at 0.4 m/s, 0.0482 (0.0137) at 0.5 m/s, and 0.0249 (0.0180) at 0.7 m/s.

Piecewise regression analysis revealed no significant association between speed and non-paretic PSR at lower speeds (slope = 0.095, p = 0.131, R^2^ = 0.06), but a significant negative association at higher speeds (slope = −0.116, p = 0.019, R^2^ = 0.38) (Figure *4*C).

## Discussion

The present study demonstrates that increased walking speed in post-stroke hemiparetic gait is associated with a reorganization of gait mechanics rather than restoration of underlying limb-level mechanical function. Although higher walking speeds were accompanied by the emergence of a structured four-phase COM power pattern characteristic of typical walking, this structural transition was not accompanied by reductions in interlimb asymmetry or improvements in propulsion–support coupling. These findings indicate that gait structure and mechanical function are partially decoupled, such that the emergence of more “normal” gait patterns reflects recovery of gait organization, but not recovery of limb-level mechanical capacity, as evidenced by persistent paretic work deficits. This study identifies a dissociation between recovery of gait organization and restoration of limb-level mechanical function, revealed through changes in COM power structure.

A central observation of this study is the presence of a transition in gait organization across walking speeds [48], occurring at approximately 0.5 m/s. At lower speeds, COM power profiles exhibited a simplified two-phase pattern, reflecting limited coordination between propulsion and support [49]. At higher walking speeds, a four-phase pattern emerged, including identifiable push-off and preload phases, indicating a qualitative reorganization toward more structured step-to-step transition mechanics [36,39]. Importantly, this transition represents a qualitative change in how mechanical work is organized across the gait cycle [34,35,47], rather than a gradual scaling of the same impaired pattern. Despite this organizational recovery, key indicators of mechanical function—notably interlimb work asymmetry and paretic propulsion relative to support—remained substantially impaired [50,51]. This dissociation demonstrates that structured gait dynamics can emerge without normalization of the limb-level mechanical deficits that generate those dynamics.

Together, these findings support a two⁳level interpretation of post⁳stroke gait recovery, in which recovery of gait organization—reflected by the emergence of structured COM power patterns[36]—can occur independently of recovery of limb⁳level mechanical capacity [30,40,50]. By jointly evaluating COM work structure and work distribution, this study provides a mechanistic basis for distinguishing organizational recovery from persistent mechanical impairment.

These findings refine the interpretation that improvements in walking speed necessarily indicate recovery [52]. In both clinical and research contexts, the emergence of identifiable push-off and multi-phase COM power patterns should be interpreted as evidence of recovery of gait organization[36,39], rather than recovery of underlying limb-level mechanical function. The present results show that such features can arise through reorganization of coordination strategies that enable higher walking performance without rectifying persistent impairments. Thus, walking speed reflects the behavioral expression of improved gait coordination, but does not directly indicate restoration of limb-level mechanical capacity [39,46,48].

The propulsion–support ratio (PSR) provides further insight into the nature of these persistent deficits. Across all walking speeds, the paretic limb exhibited a limited ability to convert vertical load support into forward propulsion, indicating reduced mechanical effectiveness. While impaired paretic propulsion is well documented in post-stroke gait [12,40,50], PSR reframes this deficit in terms of impaired coupling between support and progression. This perspective highlights that effective walking requires not only force production, but appropriate coordination of how vertical support is redirected into forward motion. The persistence of low PSR across walking speeds indicates that this coupling remains impaired even as overall gait organization improves.

From a mechanistic standpoint, these results suggest that post-stroke walking reflects a process of recovering coordinated gait organization within persistent constraints on limb-level mechanical capacity. As gait coordination improves, individuals become capable of organizing mechanical work in a more structured manner [50], reflected by the emergence of a four-phase COM power pattern and supporting higher walking speeds [48]. Importantly, this recovery of organizational capacity does not imply restoration of paretic limb mechanics [12,16]. In this framework, recovery of gait organization precedes and is partially independent of recovery of limb-level mechanical capacity: whole-body coordination may improve and enhance walking performance while mechanical deficits at the paretic limb persist.

These findings have important implications for the interpretation of gait performance. Walking speed remains a valuable descriptor of functional walking ability [11,13,14,26,28–31,44,45]; however, it provides limited information about how that performance is mechanically achieved [17,18]. The present results demonstrate that increases in walking speed and the emergence of organized gait structure reflect recovery of gait coordination [39,48], whereas persistent deficits in propulsion and interlimb work distribution indicate incomplete recovery at the limb-level mechanical domain. Incorporating biomechanically informed measures, such as COM work asymmetry and propulsion–support coupling, may therefore enable a more nuanced interpretation of walking behavior by distinguishing organizational recovery from residual mechanical impairment.

Several limitations should be considered. The sample size was modest and consisted of individuals undergoing inpatient rehabilitation, which may limit generalizability. Walking speeds were selected based on clinical tolerance rather than experimentally constrained, introducing variability while reflecting real-world conditions. The analysis was based on treadmill walking [53,54], and COM work measures require force data that may not be routinely available, although emerging measurement approaches may broaden access.

In conclusion, higher walking speeds in post-stroke hemiparetic gait are associated with a transition in gait organization characterized by the emergence of structured COM power patterns. However, this organizational recovery is not accompanied by normalization of limb-level mechanical function, as evidenced by persistent interlimb asymmetry and impaired propulsion–support coupling. These findings underscore the importance of distinguishing recovery of gait organization from recovery of mechanical capacity when interpreting walking behavior after stroke.

## Acknowledgment

This work was supported by a Natural Sciences and Engineering Research Council of Canada (NSERC) Discovery grant (04823-2017) received by J.E.A.B.

## Data Availability Declaration

The datasets used in this study are available in the author’s public repository on GitHub at: salehhosseini/Hemiparetic-dataset: Hemi Paretic Walking Dataset

## Ethical approval

Subjects provided written informed consent (Ethics ID REB21-1576)

